# A randomized controlled trial on the effects of decision aids for choosing discharge destinations of older stroke patients

**DOI:** 10.1101/2022.07.28.22277637

**Authors:** Yoriko Aoki, Kazuhiro Nakayama, Yuki Yonekura

## Abstract

In Japanese medical practice, older stroke survivors are bombarded with information regarding their discharge locations, increasing their decision-making difficulties. This study used a randomized controlled trial to evaluate the influence of using decision aids (DAs) matching the values of older stroke patients and their families on the internal conflict and participation in discharge destination decisions.

Participants were randomly allocated to intervention and control groups. The intervention lasted for two months, from admission to discharge, and a survey was conducted on both occasions. DAs were provided to the intervention group, and brochures to the control group. The decisional conflict scale (DCS) and the control preference scale (CPS) were the primary and secondary endpoints, respectively. An unpaired *t*-test and z-test analyzed inter-group differences in DCS, and CPS, respectively. This trial was registered with the University Hospital Medical Information Network (UMIN Registration No.: UMIN00032623), certified as a test registration institution by the World Health Organization.

Ninety-nine participants completed a full analysis set, which revealed that the intervention group had significantly more participants who had already decided on their discharge destination while they were admitted to the hospital. These were “the same place as before admission” in a significant number of cases. No significant inter-group differences were found in the DCS and CPS scores. DAs were effective at reducing uncertainty and controlling the decline in participation rates, especially in participants living alone who were unable to decide their discharge destination, and at clarifying the values of those aged 75 and older. The DA made it possible to increase available choices and explain the disadvantages regarding various locations of discharge destinations, allowing fewer internal conflicts in the decision-making process. Going forward, there is a need to further our understanding of methods of offering DA, the ideal duration of these interventions, and the identification of beneficiaries.

## Introduction

Medical care for strokes has advanced, and its mortality rates have drastically declined. However, age-related morbidity and recurrence rates of strokes remain high, with strokes being the second most common condition, only after dementia, leading to patients requiring long-term nursing care [1]. As a result, the roles of care personnel and the need for recovery rehabilitation enabling patients to live independently have intensified. However, individuals who have suffered a stroke experience such drastic changes in their lives that an internal conflict arises between their past self (that is hard to let go of) and their shattered self-image [2]. Due to these reasons, it is necessary to take into account numerous factors when selecting their discharge destination, including the role of their families, cognitive aspects, individual patient care behaviors and activities, health status, age, and income [3]. Patients then face a dilemma about their housing after discharge: to continue living at home or receiving care at a different location. In Japan, elders are often cared for by their families. Therefore, post-discharge housing decisions are often finalized between the family and healthcare professionals without any input from the older patients [4]. The reasons for this include difficulties in communicating with the older patients due to the severity of their condition and their families’ mindset that the older patient’s participation in decision-making is unnecessary [5]. As a result, hospitals face the challenge of coordinating among the older patients, their families, and healthcare professionals to adjust the “divergences in intentions as to discharge destination” [6]. However, there have been no established methods of aiding decision-making and no assessment criteria for decisions until now in Japan. Therefore, older patients and their families are at risk of being stricken with anxiety and remorse about the decisions made [7, 8]. The practice of shared decision-making (SDM) [9], in which patients and physicians are involved in making medical decisions together, is gradually being adopted at clinical sites. Moreover, an improved version of SDM, called the international professional SDM (IP-SDM) model [10], has now been developed that also includes families and multidisciplinary professionals in the decision-making process. This multi-professional approach has been reported as being helpful when applied to making housing decisions [11]. One method of aiding decision-making that the IP-SDM model promotes is the use of decision aids (DAs). Numerous DAs have been developed overseas and are being adopted as decision-making tools [12]. Unlike conventional informative materials, DAs compare the advantages and disadvantages of various choices and encourage choosing those that match a person’s values [13]. Some effects that have been confirmed so far and reported in all populations include increased knowledge, decreased ambiguity of internal conflicts and values, and increased participation in decision-making [14]. They have also proven to be equally effective for older people [15]. However, in the case of older people, due to reasons such as frailty and dementia, it is not easy to develop DA [16] and the progress has been limited. In Japan, patients are given informative brochures upon hospital discharge. However, the massive amounts of information in these booklets overwhelm older adults, making decision-making even more difficult [17]. However, there are no DAs in Japan that target older stroke patients or those that families and multidisciplinary professionals can use together. The likely effectiveness of such DAs is unknown.

Therefore, this study aimed to use a randomized controlled trial (RCT) to evaluate the influence of the use of DAs that match the values of older stroke patients and their families on the internal conflict over and participation in discharge destination decisions. We hypothesized that the group that is provided with a DA in selecting a discharge location will have significantly reduced decision-making conflict and increased decision-making participation compared to the non-IG. Our hypothesis was confirmed and this study was the first RCT to evaluate DA in Japan based on the values of older stroke survivors. It is hoped that the use of DAs will encourage the active participation of older stroke patients in their post-discharge housing decisions and minimize their anxiety and internal conflicts.

## Materials and methods

A protocol document for the methods of this study is included in an unpublished thesis, and we plan to publish the protocol in a journal soon.

### Study design

This study performed a two-arm parallel RCT, based on the Ottawa decision support framework [18]. This was a single-center, single-blinded test with participants allocated to the intervention group (IG) and the control group (CG) at a 1:1 ratio. The entire trial complied with CONSORT (Consolidated Standards of Reporting Trials) guidelines [19, 20], met the requirements of the CONSORT checklist, and thus conformed to the definition of a randomized test. This trial was registered with the University Hospital Medical Information Network (UMIN Registration No.: UMIN00032623), certified as a test registration institution by the World Health Organization.

### Setting

Toyama Prefectural Rehabilitation Hospital and Support Center for Children with Disabilities was made the sole institution participating in the research. The facility has 100 beds, 50 each in the third and fourth wards. The personnel who usually provide discharge assistance are physicians, nurses, physical therapists, occupational therapists, and medical social workers. While dividing roles among themselves, these multidisciplinary professionals ask older stroke patients and their families about the discharge destinations of their choice. Based on their wishes, they narrow down two to three potential facilities and social welfare services and then propose them to the patients. The staff holds numerous meetings and offers explanations orally as needed while handing out the brochures issued by the facility and municipalities. This discharge assistance method leaves the decision to multidisciplinary professionals and focuses on providing information about limited choices. Moreover, the materials offered contain vast information, such as an overview of the facilities and social welfare services. They lack content that would aid decision-making, such as the types of choices available and information on their advantages and disadvantages.

### Participants

The research participants were as follows: (1) older persons aged 65 and older, (2) those who had suffered a stroke (cerebral infarction, cerebral hemorrhage, subarachnoid hemorrhage), and (3) those admitted to rehabilitation wards during their convalescence and who had to decide their location of care after discharge. However, we excluded individuals who had difficulty making decisions because of severe dementia, aphasia, and/or an altered state of consciousness.

### Enrollment and allocation

Based on the prescribed facility criteria and preliminary survey [21], we found that the third and fourth wards were similar and concluded the baseline conditions to be the same for both in terms of patient gender, age, severity of illness, and the ratio of the number of stroke patients. About two weeks after admission, when the patients had familiarized themselves with their hospital environment, those who met the eligibility criteria were introduced to us by the head nurse. The principal investigator described the outline of the study orally to the patients, using an explanatory document. The participants were enrolled in the study after their informed consent was obtained in writing. While following the allocation table, the principal investigator randomly allocated the participants to the intervention or the CGs according to the hospital room where the initial meeting with the research participants had occurred. The principal investigator created a table by integrating (a) a random number table that Research Assistant A had created using a computer at a 1:1 ratio, and (b) an allocation table of patients according to their condition’s severity designed by the ward’s head nurse. The severity of illness was determined by the lowest total score of a daily living function assessment and the Functional Independence Measure (FIM). According to the facility criteria prescribed by the government, the severely ill are those who have a daily living function assessment of 10 points or more, or a total FIM score of 55 points or less. Until the allocation to the groups was completed, the order of allocation was concealed from Research Assistant A, the ward’s head nurse, the patients, their families, and multidisciplinary professionals (as part of the “allocation concealment mechanism”).

### The flow of selecting participants

From October 2018 to May 2020, we invited 135 individuals who had met the eligibility criteria to take part in the trial. After excluding those who had declined to take part (n = 28), we randomly allocated 107 individuals to the intervention or the CGs. Further, eight individuals were excluded with whom, in the course of follow-up, no questionnaire survey could be carried out. Finally, a total of 99 people, comprising 51 in the IG and 48 in the CG, constituted the full analysis set who were to undergo analysis (Fig 1).

**Fig 1.**
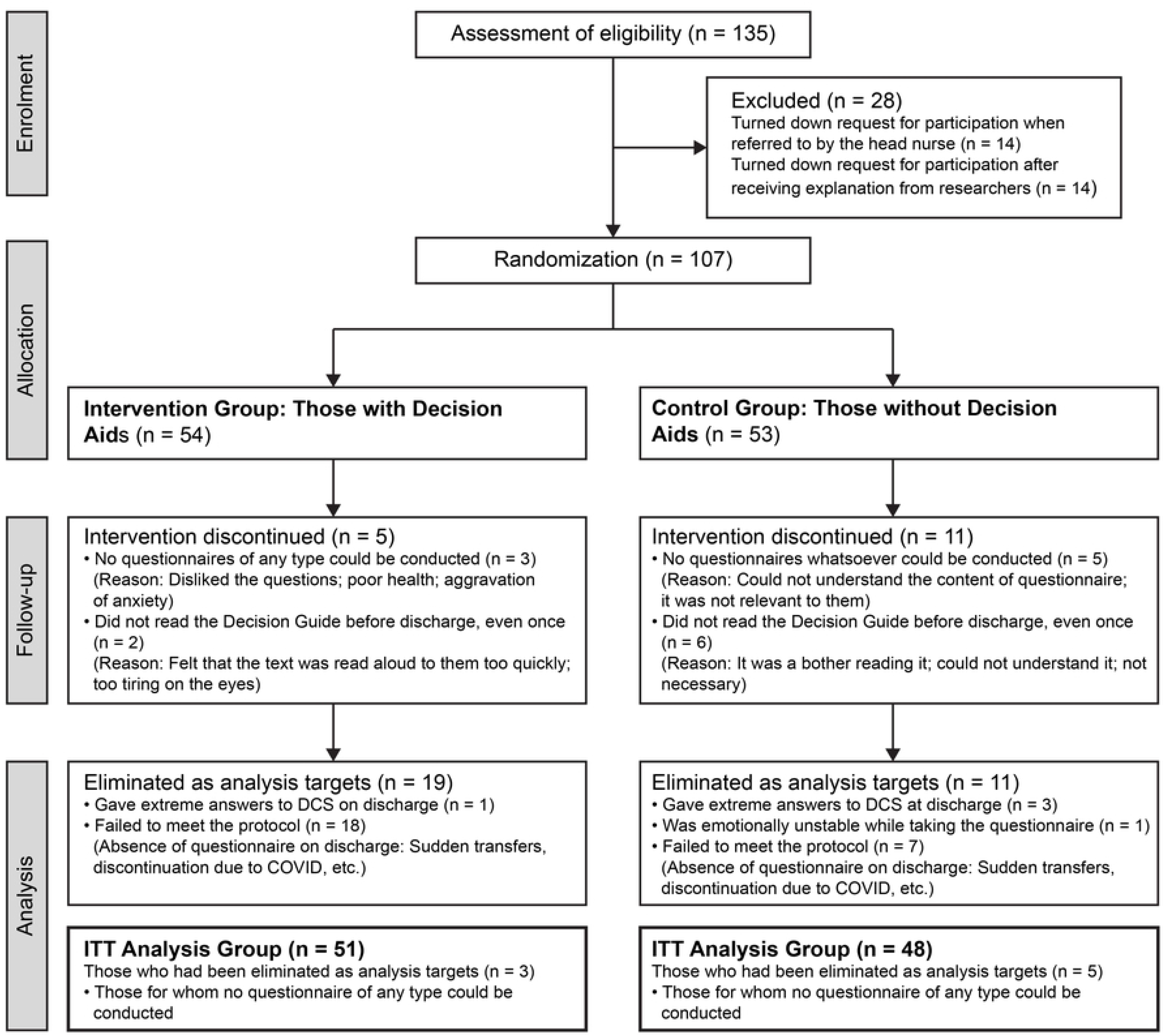
Flow diagram of the CONSORT study. The figure describes the stages of the CONSORT study, beginning with the enrollment of the participants, followed by their allocation into intervention and CGs, their follow-up, and finally the analysis of the two groups.

Although we planned to enroll 122 patients, we were not allowed to enter the hospital because of the COVID-19 pandemic; hence we halted the process temporarily. Given that the situation remained unchanged, even after a year, we decided to carry out an analysis using the number of patients we had obtained up till then.

### Intervention method

Following Coulter’s [22] systematic development process, a DA was developed based on the international patient DA standards instrument [23]. DAs consisting of 12 A4 pages were developed, listing the following six values that were common to older stroke patients and their families: (1) living standards, (2) services and costs, (3) emergencies, (4) family support, (5) environment, and (6) home repair and renovation [24].

For both groups, the duration of intervention was approximately two months, from admission to discharge. With the help of Research Assistants B (this included four research assistants who had similar roles but worked separately, as required for the study), we conducted a questionnaire survey twice, once on admission and once at discharge. The research participants and their families, the ward’s head nurse, multidisciplinary professionals, and Research Assistants B were blind to the intervention.

After a month of admission, we enquired with the IG regarding the usage of the DAs and whether they had received them. Approximately two weeks after admission, the principal investigator offered DAs to the participants in a private room. The principal investigator explained the purpose of the DA, its content, method of use, and points to note. The principal investigator ensured that the participants understood the advantages and disadvantages of the two possible post-discharge destinations, namely “the same place as before admission” and “a place different from before admission.” The investigator explained to them that the purpose of the DA was to assist them in making decisions that suited their circumstances and values. The principal investigator also explained that the content of the DA consisted of (1) information to help with devising a discharge plan, (2) information on the types and characteristics of services available, (3) information about the advantages and disadvantages of the discharge destination, (4) help with judging important values, and (5) help with organizing hospital discharge after preparations for discharge have been completed. The principal investigator explained that the patients could read the DA whenever they wished to prepare themselves for discharge and use it with their families and multidisciplinary professionals if needed. The participants were reminded not to share and show the booklets to other patients within the ward or their families by explaining to them that the efficacy of DAs has not yet been established and so two types of booklets have been handed out to all the patients to investigate their efficacy. Approximately one month after admission, after discussing the future course of action with a physician, a 15-minute interview was conducted privately to understand how the patients were using the DAs. We asked the patients if they had read the DA, used it with their family or multidisciplinary professionals, and had any questions about the content and method of using the DA after using it for approximately one month. After a month of admission, we enquired with the CG regarding the usage of the brochures (given in place of DAs) and if they had received them. The brochure’s content, describing the type and characteristics of the services available, was similar to that of the DA. The participants were explained that the brochure had been provided for their reference while deciding their discharge destinations and that it contained the same methods of usage and points of special note as those provided to the IG.

We held two meetings with the multidisciplinary professionals to explain the purpose, significance, and method of research. We explained that we could not reveal the contents of the DA or the brochure, or the allocation of patients between the two groups. We also informed them that they may respond to the queries of the older stroke patients and their families but should avoid providing instructions regarding the content of DA or about utilizing the tool. Furthermore, we asked licensed nurses and fourth-grade nursing university students who had completed their practical training to serve as Research Assistants B for conducting the questionnaire survey together. Furthermore, we trained them using a manual developed by the authors to ensure that they could provide standard and appropriate answers to anticipated questions from the research participants. (Supporting Information S1) We explained to them that we cannot reveal the allocation of the patients to them and that they were not allowed to look at the content of the DA and the brochures throughout the study duration.

### Evaluation items

The primary and secondary endpoints were evaluated, on admission and at discharge, together with the Research Assistants B, via a questionnaire survey. The primary endpoint pertained to internal conflict over decision-making and was evaluated using the 16-item Japanese-language edition of the decision conflict scale (DCS) [25]. The DCS was developed by O’Connor [26], and it is a highly reliable scale to identify the intervention effects of DAs. The test-retest reliability coefficient was 0.81, and the internal consistency coefficients ranged from 0.78 to 0.92. The Japanese-edition DCS also shows high internal consistency (Cronbach’s α: 0.84 – 0.96) [25]. DCS comprises five items, namely Sufficient explanation of information, Clarification of values, Support, Uncertainty, and Effective decision-making. Each item is evaluated using a 5-point Likert scale. All the DCS items are totaled, divided by 16, and multiplied by 25 to arrive at the total score. The total score is converted into a score ranging from 0 to 100 points, with a high score indicating a high decision-making internal conflict level. A score below 25 points indicates implementation of decision-making, and a score of 37.5 points or higher indicates a delay in decision-making and a feeling of uncertainty about its implementation [27].

The secondary endpoint pertained to participation in decision-making and was evaluated using one control preference scale (CPS) item. The CPS was developed by Strull et al. [28] and modified by Degner et al. [29]. Its reliability has been confirmed (Coombs’ criterion of 50%). The reliability of the Japanese edition of CPS has also been confirmed. The test-retest reliability of the kappa coefficient was 0.61; the weighted kappa coefficient was 0.61; and Kendall’s tau coefficient was 0.61 [30]. The role in decision-making desired by the participant is evaluated from the five written answers. Answers to Choices 1 and 2 are classified as “Active roles” (decision-making by the self), Choice 3 is classified as “Shared roles” (SDM),” and Choices 4 and 5 are classified as “Passive roles” (decision-making by others). The percentages of participation rates were also calculated, using a 10-point Visual Analog Scale.

Besides these, we asked for the following details regarding the participants’ attributes, on admission: sex, age, disease name, family makeup, the desired and the ultimate discharge destination, educational background, work history, duration of hospitalization, the status of readiness for decision-making, and the person(s) with whom a decision had been made.

### Calculation of sample size

The sample size was calculated based on the effect size of 0.3–0.4 of past studies whose primary endpoint was the DCS in the systematic review of a patient’s DAs [14]. The effect size of 0.4–0.8 shows a clinically meaningful difference in DCS and can be divided into those who make decisions and those who procrastinate [27]. Therefore, we assumed that 61 individuals were needed per group by considering a power of 0.80, an effect size of 0.5, a level of significance of two-sided α of 0.05, and losses to follow-up of 20%.

### Method of analysis

After checking the input data independently by two Research Assistants B, the primary investigator, who was not blind to the allocation process, handled the data. To retain the random allocation, we made all randomized data the targets of analysis following interventions that had been initially allocated (intention-to-treat). All the participants’ characteristics at the baseline underwent descriptive statistical testing, a *t*-test, a χ^2^-test, and a Mann–Whitney’s test. The internal conflict over decision-making, which is the primary endpoint, was subjected to an unpaired *t*-test to compare the inter-group amount of changes of the DCS subscales between the time of admission and discharge. A multiple regression analysis was also carried out to adjust the baseline values. Regarding participation in decision-making (secondary endpoint), a z-test was conducted to examine the differences in the inter-group ratios of the roles in decision-making (CPS), and a Cochran’s Q test was conducted to examine the differences in the ratio between the time of admission and discharge. An unpaired *t*-test was conducted to make inter-group comparisons between participation rates, and a paired *t*-test was conducted to compare the temporal differences in the time of admission and discharge. A subgroup analysis was also conducted on those experiencing intense internal conflict (DCS of 37.5 points or higher on admission), those living alone, older adults aged 75 and older, those who were undecided about their discharge destination in terms of the status of readiness for decision-making at their time of admission, and those experiencing long hospitalization (average duration of hospitalization: 78 days or more). SPSS Statistics for Windows, version 28 (IBM Corp., Armonk, N.Y., USA), was used for statistical analysis, and the level of significance was made two-sided, 5% or less.

## Results

### Characteristics of the participants

Table 1 summarizes the characteristics of the participants at the baseline. The participants were hospitalized for an average duration of 72.6 days (SD = 31.1) for the IG, and 82.0 days (SD = 36.3) for the CG. The average age of the participants was 75.0 years (SD = 6.4) in the IG and 75.5 years (SD = 6.6) in the CG. In both groups, a majority of the participants were males (IG = 32 [62.7%]; CG = 32 [66.7%]), many suffered cerebral infarction (IG = 36 [70.6%]; CG = 34 [70.8%]), lived with their partners (IG = 23 [46.0%]; CG = 18 [38.3%]), and had been corporate employees (IG = 29 [56.9%]; CG = 26 [54.2%]), and more than half of the participants were high school graduates or higher (IG = 40 [78.5%]; CG = 38 [79.2%]). In terms of the status of readiness for decision-making, in the IG, 66.7% had already decided on their discharge destination and 45.8% had done so in the CG. The discharge destination was the same place as before admission in 78.4% of the participants, and a different place in 21.6% of the participants. Of the 16 participants who were discharged to a place that was different from before, 6 (37.5%) were living alone. As to where the participants wanted to decide their discharge destination, the largest number of the participants wanted to do so “With their family,” followed by “With family and healthcare professionals.”

**Table 1.**
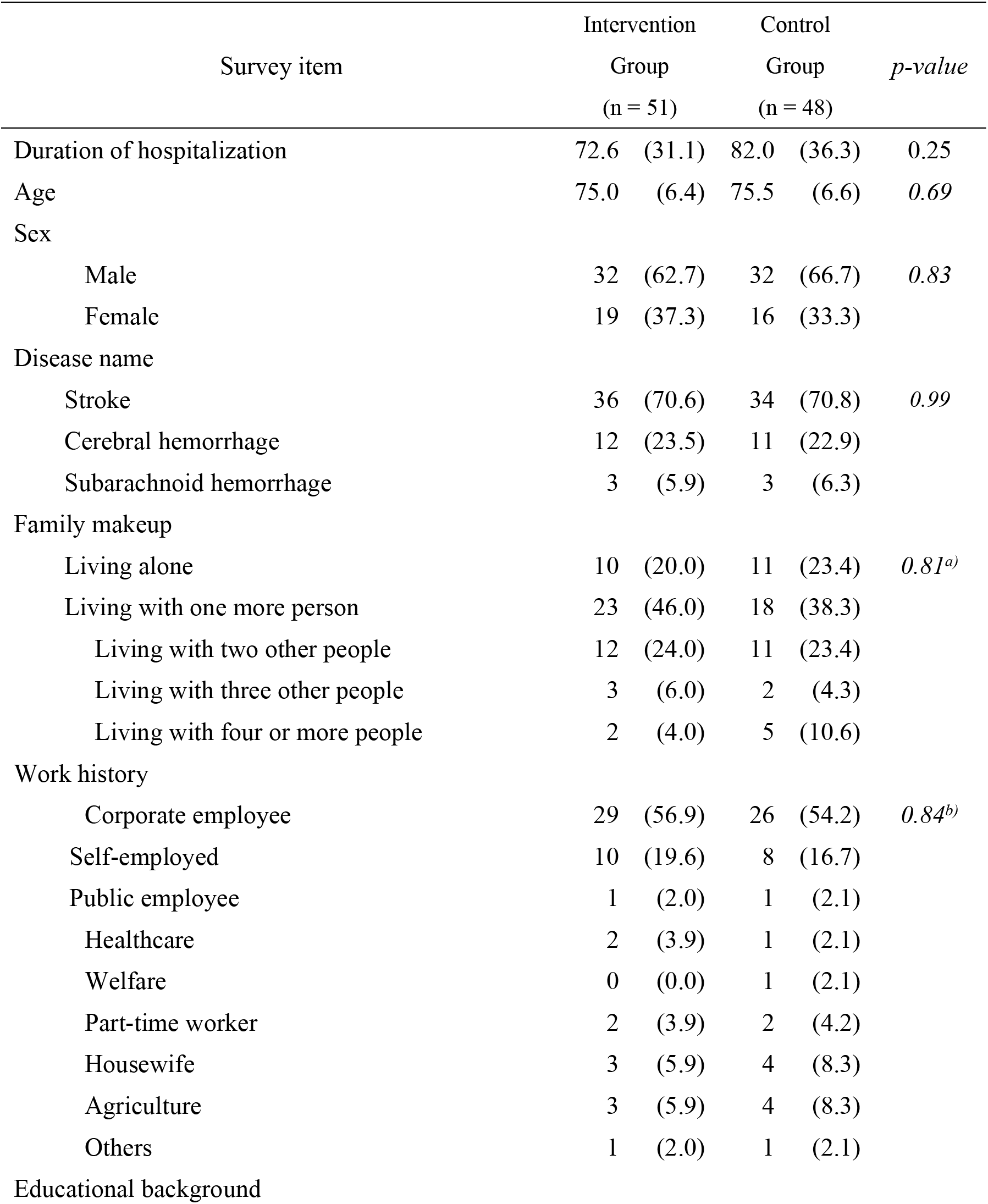
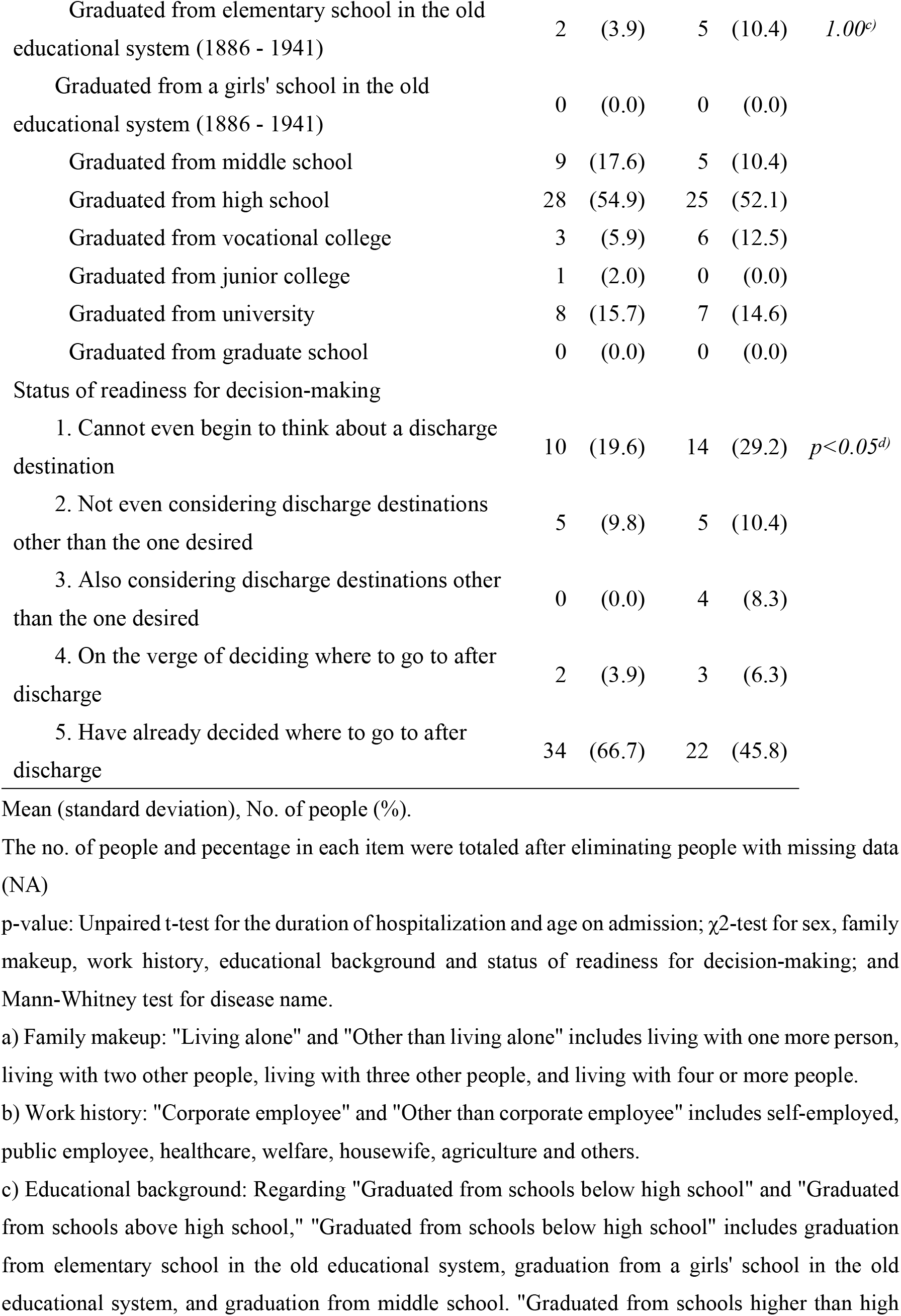
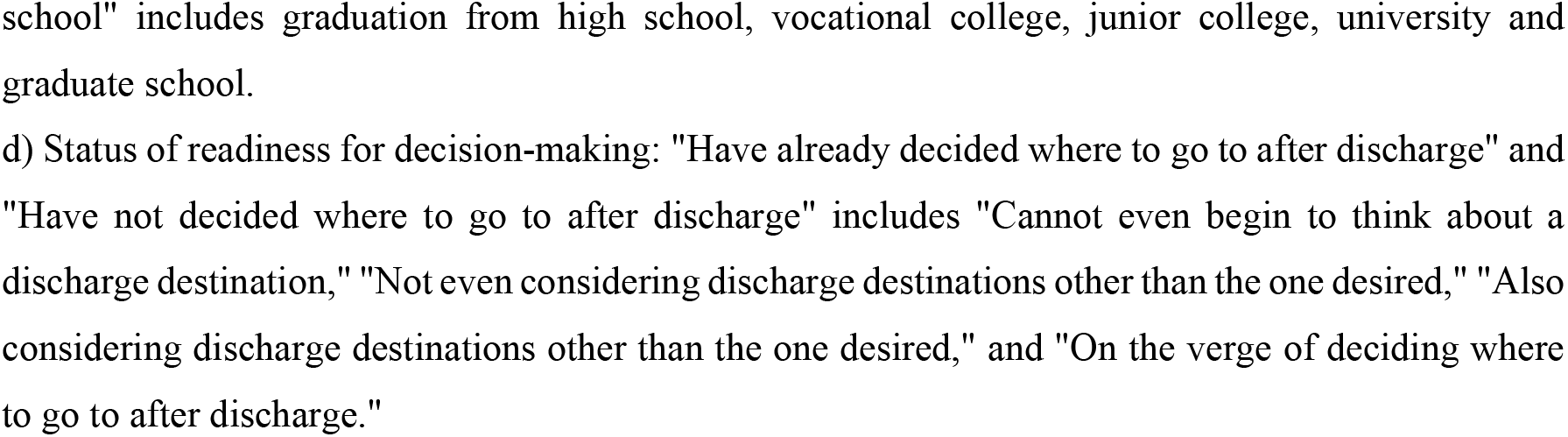
Characteristics of participants at the baseline.

We found that the IG contained significantly more participants who had already decided their discharge destination than the CG (*p* < 0.05) (Table 1). It was also found that significantly more participants chose “the same place as before admission” as their discharge destination (p < 0.01) (Table 2).

**Table 2.**
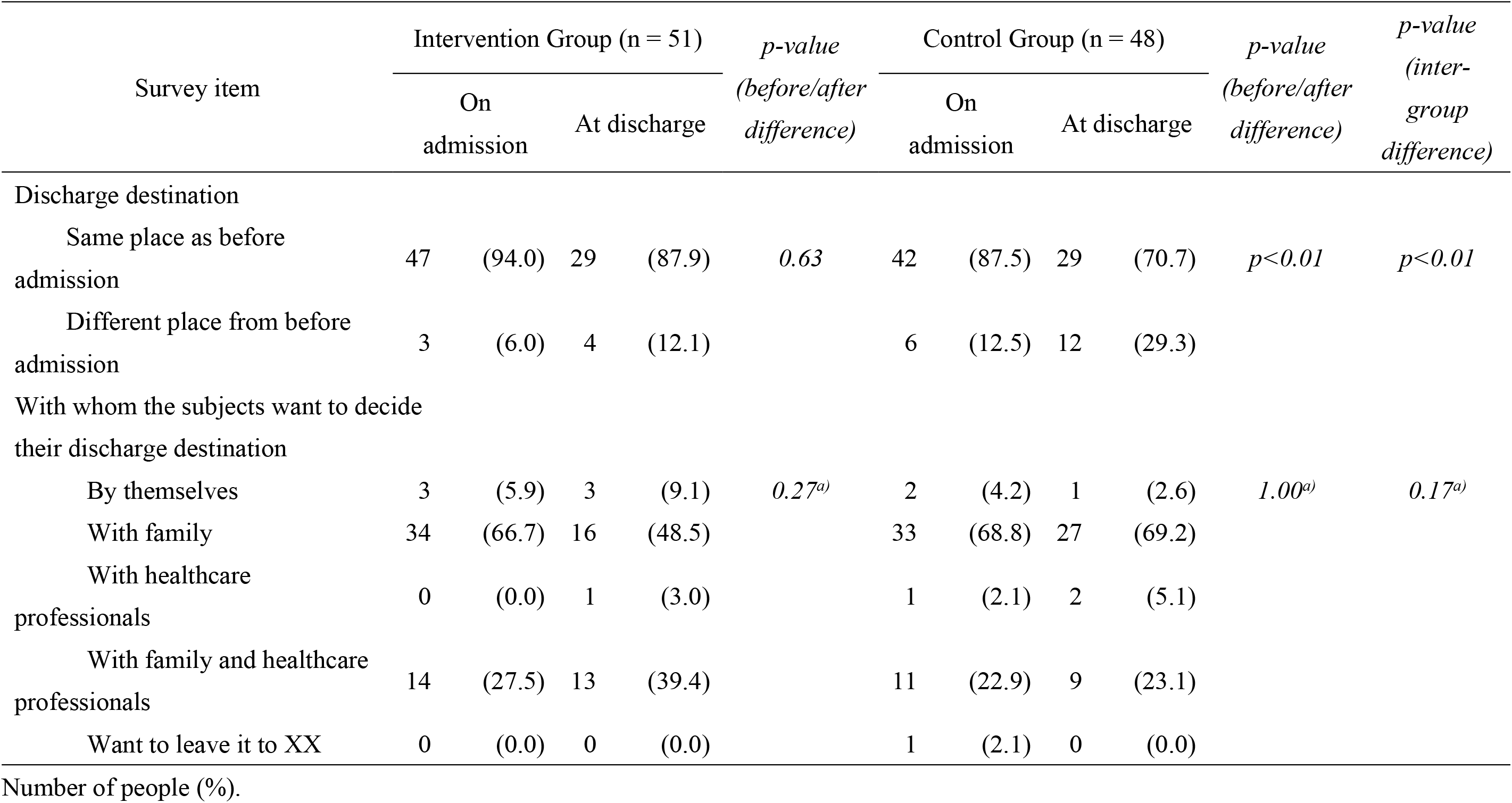
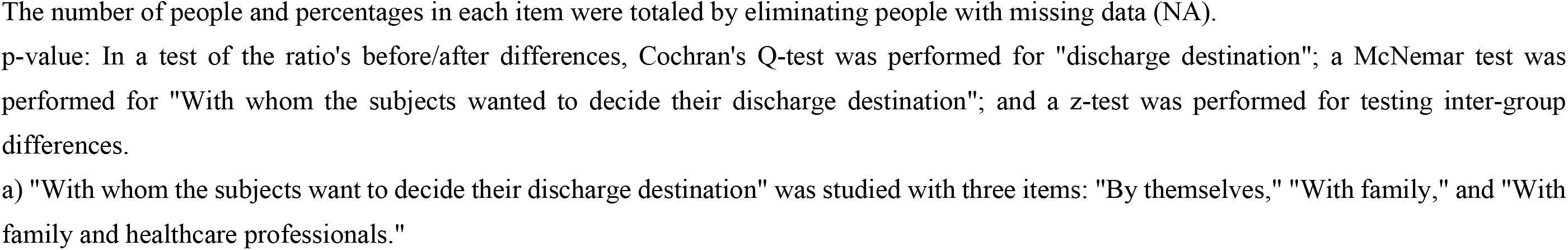
Characteristics of discharge decisions.

### Primary endpoint

In terms of conflicts over decision-making (measured by the DCS), both the intervention and CGs had intense internal conflict over “Support,” “Sufficient explanation of information,” and “Clarification of values.” The intense state of internal conflict continued even after the hospital discharge. On the contrary, the level of internal conflict over “Effective decision-making” was the lowest, during admission and discharge. No significant inter-group differences were seen in terms of the extent of change in DCS scores between admission and discharge (Table 3).

**Table 3.**
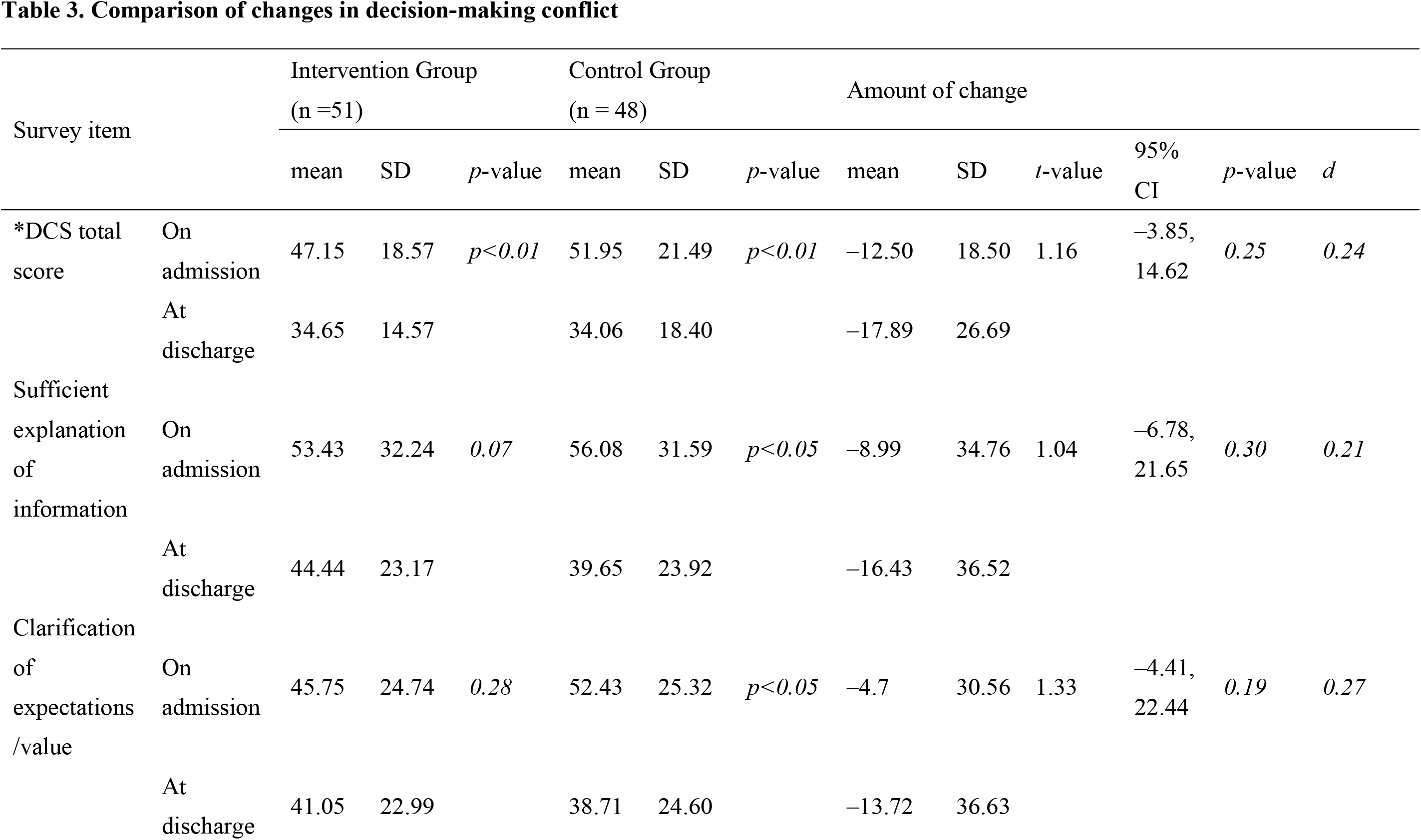
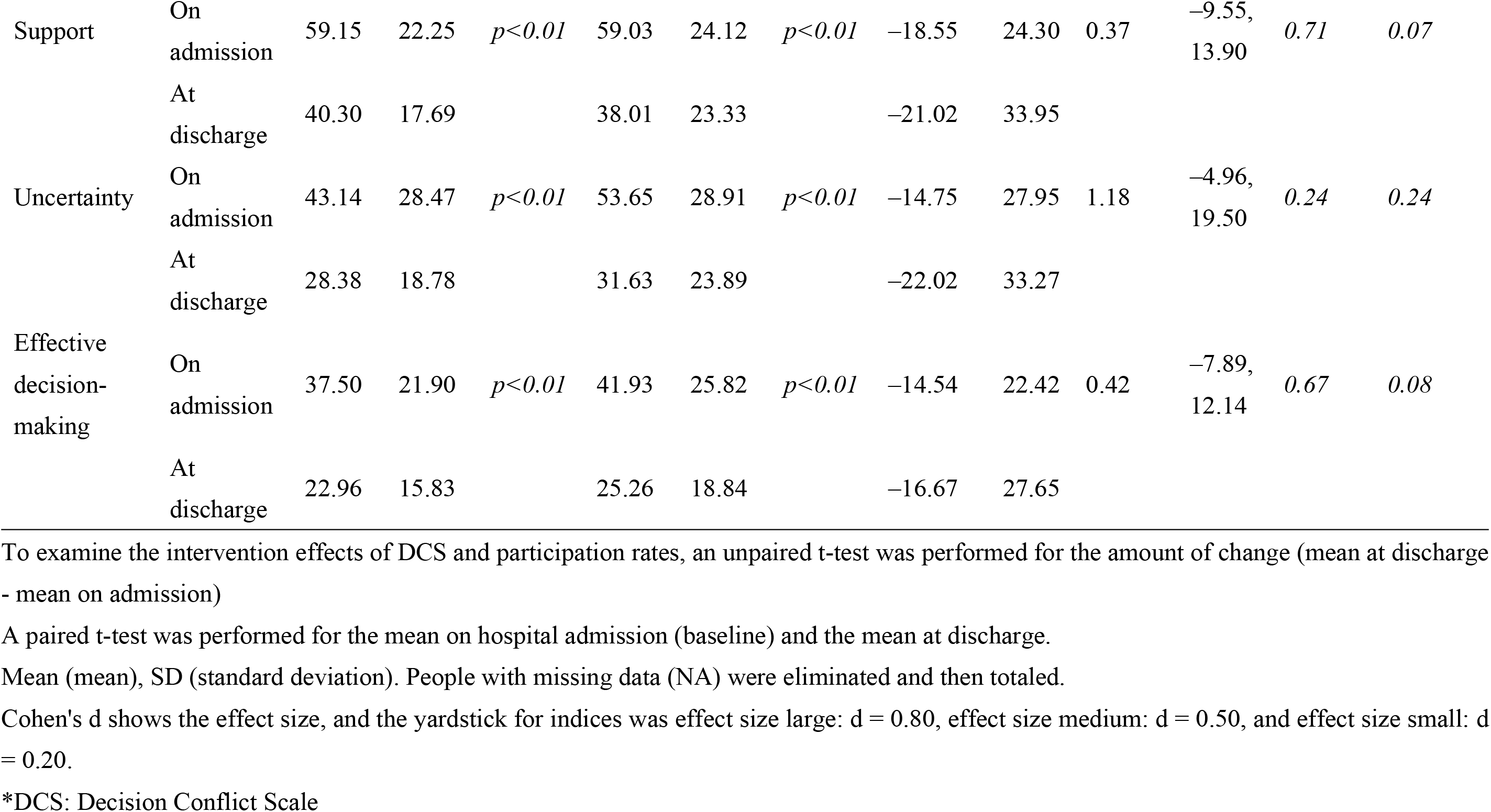
Comparison of changes in decision-making conflict.

Regarding “Uncertainty,” in particular, the number of participants who were undecided in terms of the status of readiness for decision-making showed a significantly high score *(p* < 0.05) (Table 4).

**Table 4.**
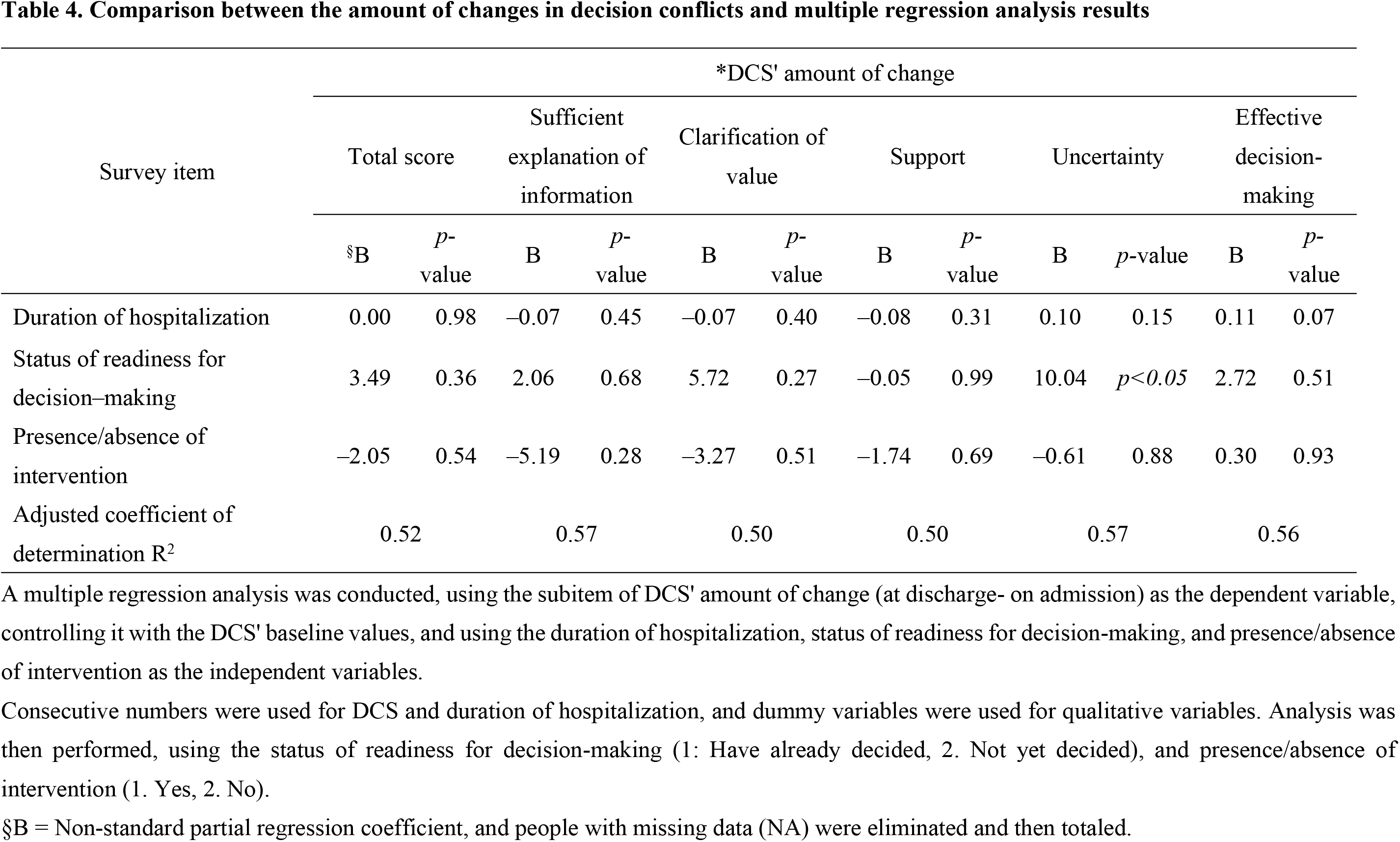
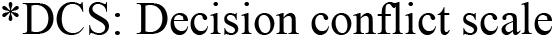
Comparison between the amount of changes in decision conflicts and multiple regression analysis results.

In terms of the effect size of the amount of change in DCS scores, a moderately significant tendency was seen with “Uncertainty” [t (21) = –1.35, p = 0.19, d = 0.59] in people who were living alone (Table 5) and with “Clarification of values” [t (49) = 1.98, p = 0.05, d = 0.57] in older adults aged 75 and older (Table 6).

**Table 5.** Difference in the means between intervention and control groups in the effects of living alone on decision conflicts.

|  | Intervention group (n=10) |  | Control group (n=11) |  | *t-value* | 95% CI | *p-value* | *d<sup>§</sup>* |
| --- | --- | --- | --- | --- | --- | --- | --- | --- |
|  | mean | SD | mean | SD |  |  |  |  |
| DCS* total score | -20.63 | 17.80 | -10.23 | 27.16 | -1.03 | -31.62, 10.82 | .32 | .45 |
| Sufficient explanation of information | -12.27 | 28.73 | -5.14 | 34.31 | -.51 | -36.21, 21.94 | .61 | .22 |
| Clarification of values | -20.04 | 30.81 | -4.42 | 33.21 | -1.11 | -44.97, 13.73 | .28 | .49 |
| Support | -17.07 | 16.01 | -20.60 | 29.12 | .34 | -18.26, 25.31 | .74 | .15 |
| Uncertainty | -31.69 | 30.78 | -11.83 | 35.89 | -1.35 | -50.55, 10.84 | .19 | .59 |
| Effective decision-making | -21.71 | 17.49 | -9.43 | 30.97 | -1.10 | -35.60, 11.03 | .28 | .48 |

An unpaired t-test was performed for amount of change (mean value at discharge- mean value on admission) to evaluate DCS intervention results among subjects living alone.
Mean and SD are shown. Calculations were made after excluding those with missing responses.
§d indicates effect size, and index criteria were as follows: large effect size: $d = .80$ , medium effect size: $d = .50$ , small effect size: $d = .20$
\*DCS: Decision Conflict Scale

**Table 6.**
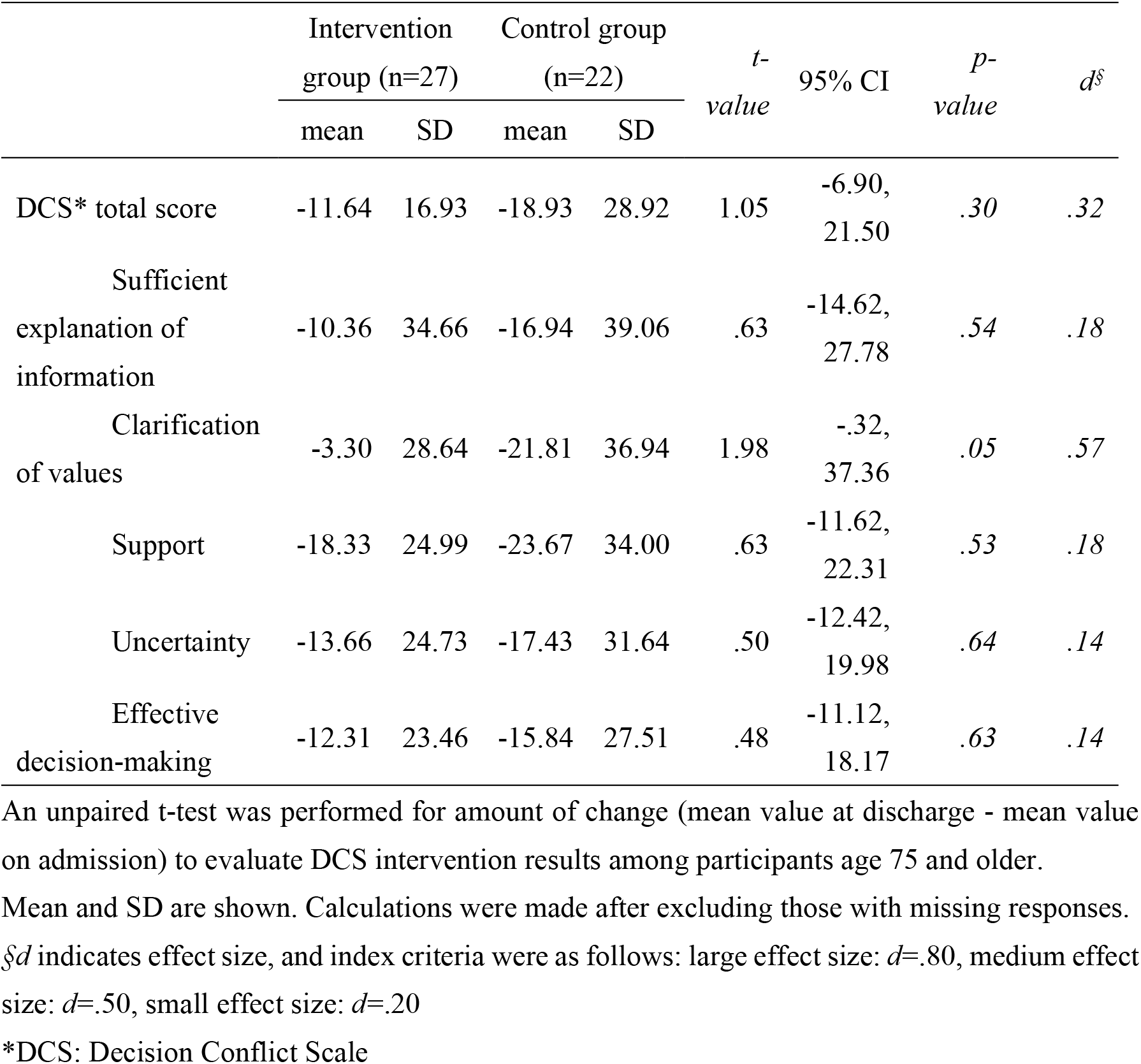
Differences in the means of intervention and control groups in decision conflicts in participants above 75.

### Secondary endpoint

In terms of participation in decision-making, as measured by the CPS, both the intervention and CGs gave the highest scores for “Active roles.” However, no significant differences were seen between the groups (Table 7).

**Table 7.** Comparison of participation in decision-making.

| Survey item | Intervention Group (*n* = 51) |  |  |  | *p*-value<br>(before<br>/after<br>differe<br>nce) | Control Group (*n* = 48) |  |  |  | *p*-value<br>(before/<br>after<br>differen<br>ce) | *p*-value<br>(inter-group<br>differe<br>nce) |
| --- | --- | --- | --- | --- | --- | --- | --- | --- | --- | --- | --- |
|  | On |  | At |  |  | On |  | At |  |  |  |
|  | admission |  | discharge |  |  | admission |  | discharge |  |  |  |
|  | n | % | n | % |  | n | % | n | % |  |  |
| *CPS |  |  |  |  |  |  |  |  |  |  |  |
| Active role | 28 | (54.9) | 19 | (57.6) |  | 27 | (56.3) | 26 | (63.4) |  |  |
| Cooperative role | 19 | (37.3) | 7 | (21.2) | *0.39* | 11 | (22.9) | 8 | (19.5) | *0.94* | *0.64* |
| Passive role | 4 | (7.8) | 7 | (21.2) |  | 10 | (20.8) | 7 | (17.1) |  |  |

*n* stands for no. of people; % shows percentages. The no. of people and percentages of each item were analyzed by eliminating people with missing data (NA).
*p*-value: Cochran's Q-test was performed to examine the before/after differences in the ratio of CPS' roles in decision-making, and a z-test was performed for testing inter-group differences.
\*CPS: Control Preference Scale

Concerning the effect size of the amount of change in participation rate, a moderately significant tendency was seen among participants living alone [t (21) = 1.44, p = 0.17, d = 0.63] (Table 8).

**Table 8.**
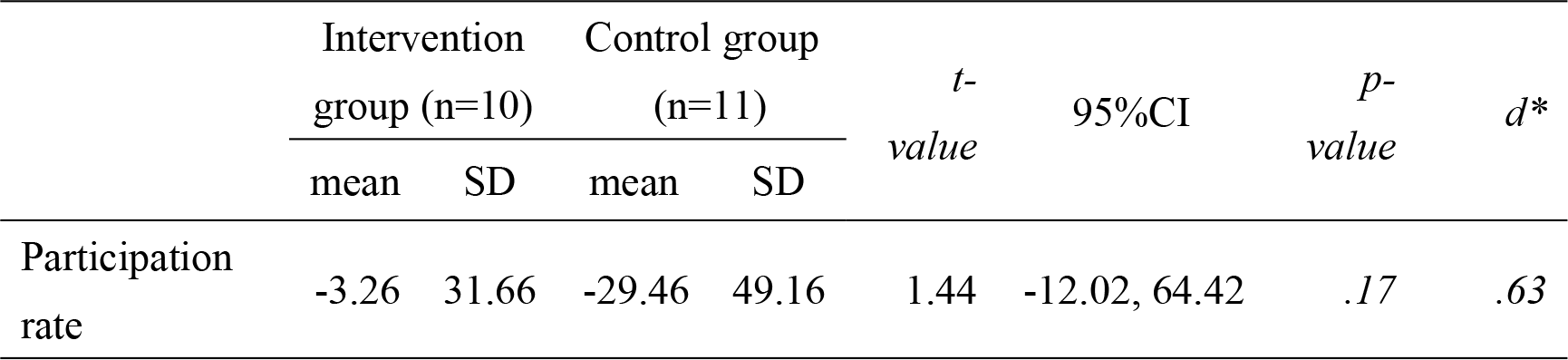
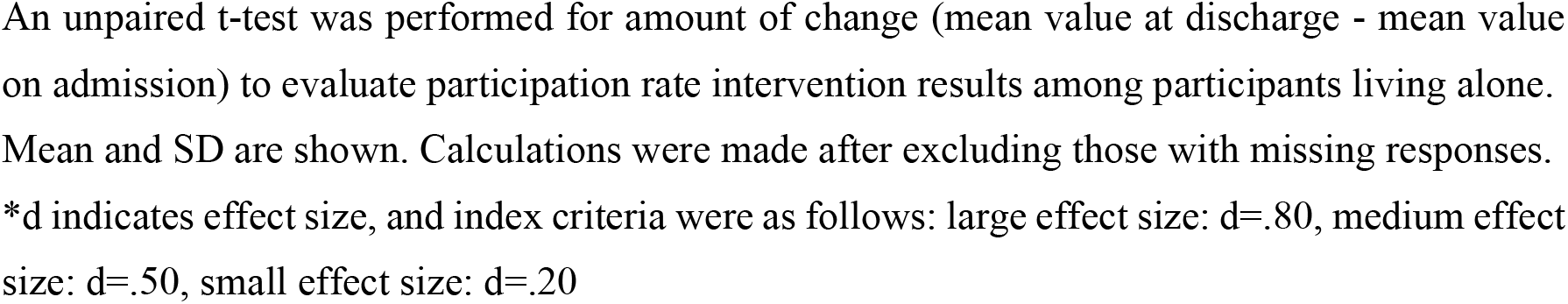
Differences in the means of intervention and control groups in the participation rates of those living alone.

## Discussion

This study examined the use of DAs based on the values held by older stroke patients and their families and used an RCT to evaluate their influence on discharge destination decisions, internal conflict, and degree of participation. Regarding internal conflict over decision-making (DCS), no significant reductions in scores were seen that were attributable to the use of DAs. A tendency to be satisfied with decision-making was observed despite high internal conflict states in Sufficient explanation of information, Clarification of values, and Support persisting at discharge, although it was not statistically significant. It has been reported that the place of convalescence desired may vary according to the participant’s condition, period, and what he/she wishes to prioritize [31]. In our study, the share of older stroke patients returning home after discharge was high—approximately 80%—which was roughly 20% higher than the share of older cancer patients [32, 33].

Moreover, older stroke patients believe, prior to hospital admission, that they would return home (which is the same place as before admission), and most felt that was the only choice available to them. Schkade and Kahneman [34] showed the tendency to use only a part of the information that may be used and underestimated information to which they do not direct their attention while making their decisions. Thus, older stroke patients who were already satisfied with being discharged to their homes may have become confused and unable to cope with the excessive information and choices they were offered. On hospital admission, the participants’ DCS scores showed high internal conflict states in all sub-items and significantly impacted the amount of change in scores from the time of discharge. This showed that the discharge destination decision caused older stroke patients’ intense internal conflict. This is reported to cause strong remorse [35] and gaps/discrepancies between the patient and their family and healthcare professionals [36]. Japan followed other countries and, in 2014, specified a DCS score of over 40 points as a condition for individuals to receive Cancer Patient Management Funding II. Assessing DCS beginning with hospital admission helps select patients who should receive nursing interventions and evaluate such nursing interventions. Our trial revealed that individuals who could not make decisions after hospital admission experienced intense internal conflict and uncertainty and that DAs reduced internal conflict caused by uncertainty, especially in people living alone. Researchers have pointed out the psychological need on the part of patients who have developed cerebrovascular disorder, a condition from which recovery is difficult to predict, and their families, to gain prospects of their home convalescent care [37]. In addition to this uncertainty of visualizing the future, of not being able to see the light at the end of the tunnel, it was believed that uncertainty would increase, in the case of those living alone, due to a shortage of support and assistance. Therefore, our study suggests the need to select people living alone as those requiring discharge assistance, from the time of hospital admission.

In our study, although “Clarification of values” indicated the smallest amount of change from admission to discharge in comparison to the other four DCS scales, it tended to reduce internal conflict in older adults aged over 75. This finding was similar to that by Stacey et al. [14] who reported little evidence that people made choices with DA that matched their values based on information (RR: 2.06, 95% CI: 1.46 – 2.91). Concerning the place of convalescence for older adults, their final abode must also be considered and the grounds for determining the best place for older adults have not yet been clarified [38, 39]. As a result, it has been revealed that diverse values exist when deciding the place of convalescence for older adults [16,40,41]. The DA used in our study was developed based on the values of older stroke patients and their families who had to choose where to live after discharge. However, all the values extracted were important, suggesting it to be difficult to differentiate them. Older adults make decisions by relying on their past experiences and predictions, making them liable to biases [42]. Older adult patients agree to return home upon being suggested so, and professionals providing them information feel no need to make an effort to describe other potential locations to the patients. This finding suggests a risk that the advantages and, especially disadvantages, cannot be compared—which also happens to be part of the decision-making process—and that values are less liable to be clarified. Dugas et al. [43] stated that involving the immediate parties in the development process helps to avoid stigma and to clarify society’s essential problems. It is also reported that DA can reduce the percentage of patients who are unable to make decisions [44]. DA was shown to have the potential to help reduce the ambiguity and uncertainty of values held, especially, by older adults aged 75 and older living alone who, despite having ample experience, are inevitably entangled in a situation in which they are affected by their surroundings and the people around them. Hence, as the result of this study suggests, DA makes it easier for older adults to decide.

Next, in terms of participation in decision-making, no significant increases were seen in the CPS scores after using DA. However, there was a tendency for DA use to control the decrease in participation rate among participants who lived alone. Our study’s percentage of CPS playing an “active role” was about 30% lower than that seen in past research of other countries [45]. Instead, the percentages were characteristically high in terms of “Cooperative role” and “Passive role,” that is, working together with other people or leaving the decision to others. However, Almborg et al. [46] report that almost none of the patients who take part in discharge planning believe that they were taking part in planning their treatment and care needs, services, rehabilitation, or goal-setting. As a result, the roles of decision-making, as evaluated by CPS, were based solely on self-reporting by older stroke patients. Hence, we feel that they have not been able to appropriately grasp whether or not they had actually participated in decision-making. As cultural characteristics of decision-making among the Japanese, Kawai et al. [47] state that the people tend to emphasize harmony, deliberately refrain from stating their opinions, leave decision-making entirely to others, and provide tacit consent. However, the fact that older stroke patients had wished to decide their discharge destination, and had acknowledged that they had taken part in them, was a new insight we gained. In our study, those who had made decisions with someone else, such as family and healthcare professionals, accounted for approximately 80% and almost no one made decisions on his/her own. As seen, even if the decisions were about older adults and they had to decide where to discharge themselves, the fact that they had decided together with family and healthcare professionals may have led to their high level of awareness that they were also taking part in the process. It has been shown that the ability to take part in decision-making (as evaluated by CPS) is influenced most strongly by a shortage of knowledge of the choices available, the patients’ preferences, and a lack of balance in power relationships [48]. Older stroke patients, expecting to return home after discharge, may have hesitated to make a decision, out of a sense of guilt and awareness of having been afflicted by a stroke and that they would therefore be highly dependent on someone else. Thus, it was suggested that DA might benefit decision-making among people who live alone and are likely to lack support. This is also the reason for the need for objective evaluations by family and healthcare professionals.

### Limitations

The DA utilized in this study was the first tool of its kind in Japan that was evaluated via an RCT targeting older stroke patients. However, it is necessary to consider several limitations while interpreting the results. This study initially verified the genuine effects only of DA, so the intervention content consisted only of the distribution of DA or brochures, confirming their usage status. Therefore, although DAs are designed to promote SDM, offering them itself does not guarantee the implementation of SDM with family and various professionals. Moreover, the difference in effects was not particularly evident because due to COVID-19 restrictions, the intended sample size could not be achieved. It is also necessary to bear in mind that the brochure’s content was the same as certain sections of the DA, and the risk of contamination would have been caused by moving people to different hospital rooms. Only one institution was used in this study as the research target facility, and there is the possibility that it has numerous unique facility criteria and regional characteristics which may not be generalizable to other institutions. Going forward, there is a need to increase research target facilities and study participants to generalize and standardize the findings and data and to further understand the period and method of offering DA as well as the selection and content of target individuals.

## Conclusion

Our study showed that DA was effective in easing the uncertainty and controlling the decline in participation rates, especially felt by people living alone who had been unable to decide their discharge destinations since the time of their hospital admission, and in clarifying the values of older people aged 75 and older. Henceforth, it is necessary to widen the choices offered to participants while taking the time to ask them about the post-discharge life they were envisaging. Then, while making use of DA, we felt that, by adding explanations of the disadvantages of the choice made, the participants could take part in decision-making, which could reduce internal conflict.

## Data Availability

Data supporting the results reported in the article are available upon request from the principal investigator's affiliation, Dept. of Gerontological Nursing, Faculty of Medicine, University of Toyama. Yoriko Aoki, +81-74-434-7420.

## Acknowledgments

We would like to thank all participating older stroke patients and their families, as well as the staff of the Toyama Prefectural Rehabilitation Hospital & Support Center for Children with Disabilities.

## Supporting Information

**S1 File.** Manual for Research Assistants

## References

1. Ministry of Health, Labour and Welfare. White paper on aging society. 2021 ed. (entire edition). Available from: https://www8.cao.go.jp/kourei/whitepaper/w-2021/zenbun/pdf/1s2s_02.pdf

2. Ojima K, Ito M, Yajima M. The psychological process in the recovery phase of latter-stage elderly who became hemiplegic because of a cerebrovascular disease. The Bull Sci Nurs Res Ashikaga Univ. 2019;7: 1–12.

3. Roy N, Dubé R, Després C, Freitas A, Légaré F. Choosing between staying at home or moving: A systematic review of factors influencing housing decisions among frail older adults. PLOS ONE. 2018;13: e0189266. doi: 10.1371/journal.pone.0189266.

4. Ono M, Fukuda H, Otsu S, Uchida M, Awaya N. Care to encourage participation of the older adult in need of long-term care in the post-discharge decision-making process: An analysis of responses shown to families and medical staff, Japanese. J Clin Nurs. 2006;32(2): 266–271.

5. Iso R, Iijima S. The current status and challenges of a client’s participation and decisions in multidisciplinary and inter-institution cooperation at the time of elderly’s hospital discharge. The Journal of the International University of Health and Welfare. 2016;21: 10–20.

6. Harada K, Matsuda C, Nagahata T. An acute-care hospital’s discharge adjustments: Difficulties in assisting the discharge of the elderly as perceived by nurses. J Jpn Acad Gerontol Nurs. 2014;18: 67–75.

7. Okumura Y, Yokoi S, Hashimura H, Takigawa K. Anxiety felt by main caretakers of cerebrovascular disease patients admitted to convalescence rehabilitation wards: Changes between about one week after admission and before discharge. J Nurs. Shiga University of Medical Science. 2012;10(1): 34–37.

8. Sakai K, Tsukahara C, Iwaki N, Makino T. Factors of patients and families that affect the decision on the location of care for progressive cancer patients. Ishikawa J Nurs. 2011;8: 41-50.

9. Kon AA. The shared decision-making continuum. JAMA. 2010;304: 903–904. doi: 10.1001/jama.2010.1208.

10. Légaré F, Adekpedjou R, Stacey D, Turcotte S, Kryworuchko J, Graham ID, et al. Interventions for increasing the use of shared decision making by healthcare professionals. Cochrane Database Syst Rev. 2018;7: CD006732. doi: 10.1002/14651858.CD006732.pub4.

11. Légaré F, Stacey D, Brière N, Robitaille H, Lord MC, Desroches S, et al. An interprofessional approach to shared decision making: An exploratory case study with family caregivers of one IP home care team. BMC Geriatr. 2014;14: 83. Available from: http://www.biomedcentral.com/1471-2318/14/83. doi: 10.1186/1471-2318-14-83.

12. A to Z inventory of decision Aids; 2019. Alphabetical List of Decision Aids by Health Topic. Available from: https://decisionaid.ohri.ca/AZlist.html. The Ottawa Hospital Research Institute

13. O’Connor AM, Stacey D, Jacobsen MJ. Ottawa Personal Decision Guide for People Facing Tough Health or Social Decisions. Ottawa Hospital Research Institute & Canada: University of Ottawa. 2012 [updated 2015]. Available from: https://decisionaid.ohri.ca/docs/das/OPDG.pdf.

14. Stacey D, Légaré F, Lewis K, Barry MJ, Bennett CL, Eden KB, et al. Decision aids for people facing health treatment or screening decisions. Review. Cochrane Database Syst Rev. 2017; 4: CD001431. doi: 10.1002/14651858.CD001431.pub5.

15. van Weert JC, van Munster BC, Sanders R, Spijker R, Hooft L, Jansen J. Decision aids to help older people make health decisions: A systematic review and meta-analysis. BMC Med Inform Decis Mak. 2016;16: 45. doi: 10.1186/s12911-016-0281-8.

16. Garvelink MM, Emond J, Menear M, Brière N, Freitas A, Boland L, et al. Development of a decision guide to support the elderly in decision making about location of care: An iterative, user-centered design. Res Involv Engagem. 2016;2: 26. doi: 10.1186/s40900-016-0040-0.

17. Nakayama K, Setoyama Y, Fujii T, Shinozaki E, Aida T, Takaki H, et al. Chapter 4. Nursing and information, systematic nursing course – Separate volume – Nursing informatics. Tokyo: Igaku Shoin; 2017. pp. 63-88.

18. Jacobsen MJ, O’Connor AM, Stacey D. Decisional needs assessment in populations. 1999 [updated 2013; cited 2022 Mar 8]. Available from: https://decisionaid.ohri.ca/eTraining/docs/s6_Population_Needs_Assessment.pdf

19. Moher D, Schulz KF, Altman DG. The CONSORT statement: Revised recommendations for improving the quality of reports of parallel-group randomised trials. Lancet. 2001;357: 1191–1194. doi: 10.1016/S0140-6736(00)04337-3.

20. Tsuya K, Kamioka H, Origasa H, Sato H. Introduction and explanation of the 2017 CONSORT non-drug intervention edition. 2017 CONSORT NPT Extension. Jpn Pharmacol Ther. 2019;47: 865-884.

21. Aoki Y, Nakayama K. Factors that affect elderly stroke patients’ discharge destinations in convalescence rehabilitation wards. J Int Nurs Res. 2019;42: 881–888.

22. Coulter A, Stilwell D, Kryworuchko J, Mullen PD, Ng CJ, van der Weijden T. A systematic development process for patient decision aids. BMC Med Inform Decis Mak. 2013;13(Suppl 2): S2. Available from: http://www.biomedcentral.com/1472-6947/13/S2/S2. doi: 10.1186/1472-6947-13-S2-S2

23. Elwyn G, O’Connor AM, Bennett C, Newcombe RG, Politi M, Durand MA, et al. Assessing the quality of decision support technologies using the International Patient Decision Aid Standards instrument (IPDASi). PLOS ONE. 2009;4: e4705. doi: 10.1371/journal.pone.0004705.

24. Aoki Y. The development process of a “Guidebook for considering discharge destination” for elderly patients hospitalized due to stroke, and challenges for promoting its widespread use. J St Lukes Soc Nurs Res. 2022;25: 52–54.

25. Kawaguchi T, Azuma K, Yamaguchi T, Soeda H, Sekine Y, Koinuma M, et al. Development and validation of the Japanese version of the Decisional Conflict Scale to investigate the value of pharmacists’ information: A before and after study. BMC Med Inform Decis Mak. 2013;13: 50. doi: 10.1186/1472-6947-13-50.

26. O’Connor AM. Validation of a decisional conflict scale. Med Decis Making. 1995;15: 25–30. doi: 10.1177/0272989X9501500105.

27. O’Connor AM. User Manual-Decision Conflict Scale. 1993 [updated 2010]. Available from: www.ohri.ca/decisionaid

28. Strull WM, Lo B, Charles G. Do patients want to participate in medical decision making? JAMA. 1984;252: 2990–2994. doi: 10.1001/jama.1984.03350210038026.

29. Degner LF, Sloan JA, Venkatesh P. The control preferences scale. Can J Nurs Res. 1997;29: 21–43.

30. Azuma K, Kawaguchi T, Yamaguchi T, Motegi S, Yamada K, Onda K, et al. Development of Japanese versions of the control preferences scale and information needs questionnaire: Role of decision-making and information needs for Japanese breast cancer patients. Patient Prefer Adherence. 2021;15: 1017–1026. doi: 10.2147/PPA.S295383.

31. A committee to investigate the raising of public awareness of medical care in the final stages of life. A report on the awareness survey regarding medical care at the final stage of life. 2018 [cited 2022 Mar 8]. Available from: https://www.mhlw.go.jp/toukei/list/dl/saisyuiryo_a_h29.pdf

32. Kojima K, Shiraishi S. Investigation of factors relating to stroke rehabilitation patients’ discharge to home: Utilizing the data of rehabilitation patient databank. J Health Sci. Nihon Fukushi University. 2015;18: 9–17.

33. Omachi I, Yokoo S, Tamura H, Nozoe A, Matsufuji Y, Morizono K, et al. Factors related to elderly cancer patients’ switching from hospitals to home care. Bull Kyushu Agric Exp Stn. 2013;22: 9-13.

34. Schkade DA, Kahneman D. Does living in California make people happy? A Focusing Illusion in Judgments of Life Satisfaction. Psychol Sci. 1998;9: 340–346. doi: 10.1111/1467-9280.00066.

35. Elidor H, Ben Charif A, Djade CD, Adekpedjou R, Légaré F. Decision regret among informal caregivers making housing decisions for older adults with cognitive impairment: A cross-sectional analysis. Med Decis Making. 2020;40: 416–427. doi: 10.1177/0272989X20925368.

36. O’Connor AM, Jacobsen MJ. Workbook on developing and evaluating decision Aids. 2003 [cited 2022]. Available from: www.ohri.ca/decisionaid

37. Kajiya M, Moriyama M. Psychological care needs of cerebrovascular disorder patients and their families, three months after onset. The Japanese Journal of Research in Family Nursing. 2010;16: 71-80.

38. Boland L, Légaré F, Perez MM, Menear M, Garvelink MM, McIsaac DI, et al. Impact of home care versus alternative locations of care on elder health outcomes: An overview of systematic reviews. BMC Geriatr. 2017;17: 20. doi: 10.1186/s12877-016-0395-y.

39. Leeds L, Meara J, Hobson P. The impact of discharge to a care home on longer term stroke outcomes. Clin Rehabil. 2004;18: 924–928. doi: 10.1191/0269215504cr807oa.

40. Murray MA. When you need extra care, should you receive it at home or in a facility? Ottawa Patient Decision Aid Research Group. 2010. Available from: https://decisionaid.ohri.ca/docs/das/Place_of_Care.pdf

41. Shimpo H. Social work on discharge assistance: Formation of “situational values” in the patient support system. Tokyo: Aikawa Books; 2014. p. 68.

42. Otake F, Hirai T. Behavioral economics at the sites of medicine: Doctors and patients missing each other. Tokyo: Toyo Keizai; 2018. p. 166.

43. Dugas M, Trottier MÈ, Chipenda Dansokho SC, Vaisson G, Provencher T, Colquhoun H, et al. Involving members of vulnerable populations in the development of patient decision aids: A mixed methods sequential explanatory study. BMC Med Inform Decis Mak. 2017;17: 12. doi: 10.1186/s12911-016-0399-8.

44. Hoefel L, O’Connor AM, Lewis KB, Boland L, Sikora L, Hu J, et al. 20th anniversary update of the Ottawa Decision Support Framework Part 1: A systematic review of the decisional needs of people making health or social decisions. Med Decis Making. 2020;40: 555–581. doi: 10.1177/0272989X20936209. Appendix B.

45. Adekpedjou R, Stacey D, Brière N, Freitas A, Garvelink MM, Turcotte S, et al. “Please listen to me”: A cross-sectional study of experiences of seniors and their caregivers making housing decisions. PLOS ONE. 2018;13: e0202975. doi: 10.1371/journal.pone.0202975.

46. Almborg AH, Ulander K, Thulin A, Berg S. Discharge planning of Stroke Patients: The Relatives’ Perceptions of participation. J Clin Nurs. 2009;18: 857–865. doi: 10.1111/j.1365-2702.2008.02600.x.

47. Kawai N, Sugaya A, Morino A, Imaizumi K, Yanaida K, Sakai S, et al. Japan’s cultural characteristics in healthcare mentioned in overseas literature. J Chiba Acad Nurs Sci. 2007;13: 119–127.

48. Joseph-Williams N, Elwyn G, Edwards A. Knowledge is not power for patients: A systematic review and thematic synthesis of patient-reported barriers and facilitators to shared decision making. Patient Education and Counseling. Patient ed. 2014;94: 291–309. doi: 10.1016/j.pec.2013.10.031.

